# The modified arterial reservoir: an update with consideration of asymptotic pressure (*P*_∞_) and zero-flow pressure (*P*_*zf*_)

**DOI:** 10.1101/2020.01.22.20018440

**Authors:** Alun D Hughes, Kim H Parker

## Abstract

This article describes the modified arterial reservoir in detail. The modified arterial reservoir makes explicit the wave nature of both reservoir (*P*_*res*_) and excess pressure (*P*_*xs*_). The mathematical derivation and methods for estimating *P*_*res*_ in the absence of flow velocity data are described. There is also discussion of zero-flow pressure (*P*_*zf*_), the pressure at which flow through the circulation ceases; its relationship to asymptotic pressure (*P*_*∞*_) estimated by the reservoir model; and the physiological interpretation of *P*_*zf*_. A systematic review and meta-analysis provides evidence that *P*_*zf*_ differs from mean circulatory filling pressure.

## Introduction

The concept of an arterial reservoir dates back to Borelli(1) and Hales(2); it was developed further by Weber (from whom the term Windkessel is derived(3)(cited in(4))), and by Frank who provided a mathematical framework for it.(5) More recently in the early 2000’s a revised form of the arterial reservoir was proposed by Parker and Tyberg(6) and results using this approach were first published by Wang et al.(7) According to this model the pressure waveform was envisaged as the sum of a Windkessel pressure and (mainly forward) travelling waves.(7) While this proposal elicited interest, it also received criticism,(8-11) with criticisms related to the assumption of a uniform Windkessel pressure being particularly pertinent. More recently the model was revised to address this problem.(12) In the revised model the Windkessel pressure was replaced by a reservoir pressure, which was made up from waves and was delayed by the time taken for waves to travel from the aortic root to the location of measurement.(12) This modification makes explicit the wave nature of reservoir pressure and this modified definition has achieved some degree of acceptance.(13) The aim of this review is to describe the modified arterial reservoir in more detail, and to provide more information regarding the asymptotic pressure, *P*_*∞*,_ its relationship to zero-flow pressure (*P*_*zf*_), the arterial pressure at which flow through the circulation ceases, also termed critical closing pressure,(14)) and the physiological interpretation of *P*_*zf*_.

## Wave travel in arteries and its relation to the reservoir

The existence of wave travel in arteries is undisputed.(13) A number of studies have envisaged the arterial system as a single or a T-tube; however this approach to arterial hemodynamics is too simplistic and a more sophisticated model of wave propagation in arteries is necessary.(15) More realistic 1-dimensional models show that the branching pattern of the arterial circulation gives rise to myriad reflected waves, which are themselves re-reflected and re-re-reflected before returning to the aortic root.(16, 17) Tapering of the arterial system may also make an important contribution to wave reflection patterns,(18) and inclusion of visco-elastic behaviour may also be important in intermediate size vessels.(19) The reservoir pressure can be understood as the pressure due to the cumulative effect of these reflected and re-reflected waves, which decrease in magnitude but increase in number as they travel. Another implication of the branching nature of the arterial tree is that the reflection coefficient at a bifurcation depends on the direction of travel of the wave. At bifurcations in large arteries the combined admittance of the offspring arteries is similar to that of the parent artery (i.e. most bifurcations are well-matched for forward travelling waves); however this also means they are poorly-matched for backward travelling waves.(17, 20) Thus, large reflections from peripheral reflection sites, are dispersed by the re-reflections they undergo while travelling back to the aortic root. These considerations account for a ‘horizon effect’ where the apparent time of reflection of the initial compression wave, as indicated by a peak in the backward wave intensity, is independent of the site where the measurements are made.(16)

## A modified definition of reservoir pressure

The modified definition of the reservoir pressure (*P*_*res*_) assumes that the reservoir is made up of a network of *N* arteries. It is also assumed that the root artery (the aorta), *A*_*0*_, is connected to the left ventricle and receives the stroke volume, *Q*_*in*_, and that there are *K* terminal vessels – these are assumed to be connected to the microcirculation which is not considered part of the reservoir. A time-varying average pressure in each vessel *P(t, n)* is defined as the integral of pressure over the length of each arterial segment. A simplified version of this scheme is illustrated in Figure 1.

**Figure 1.**
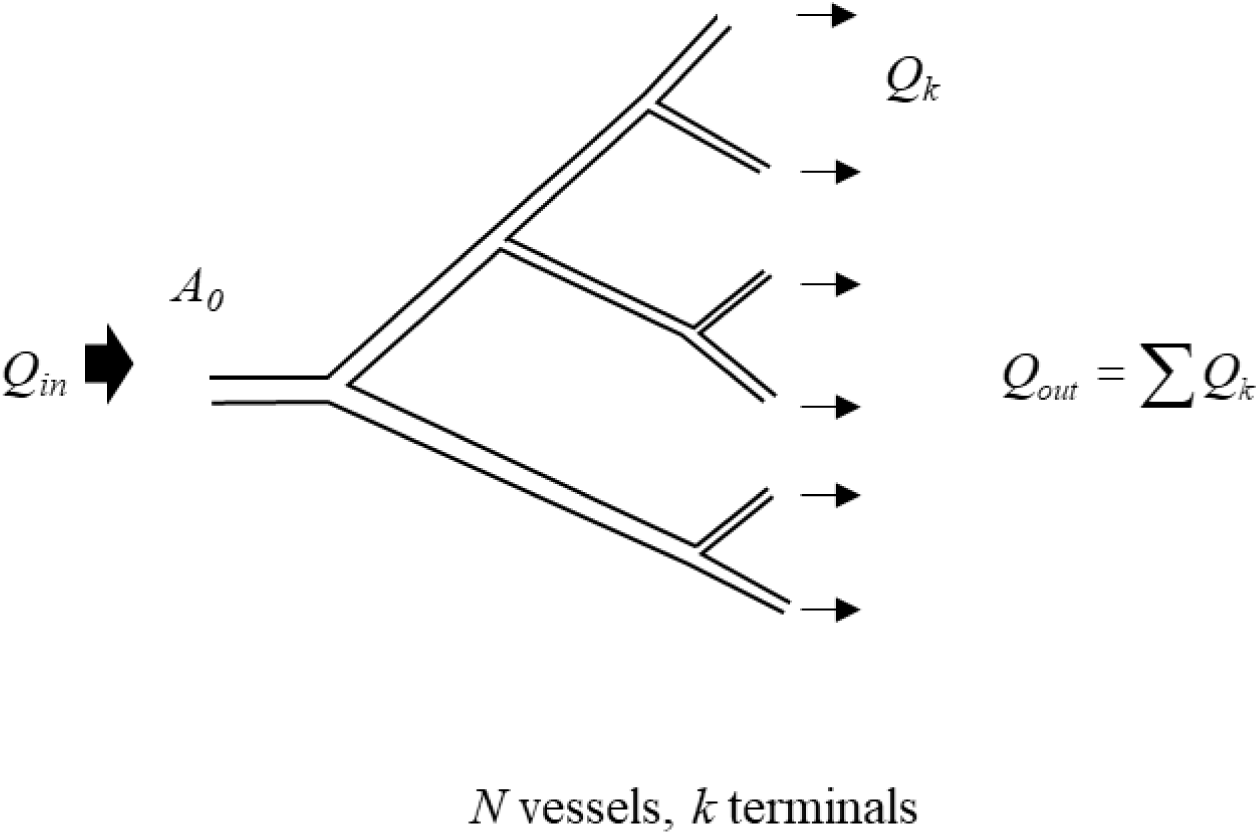
A simplified schematic showing a network of branching arteries corresponding to an arterial reservoir (where *N*=*11* and *K*=*6*). The inlet to the network, A_0_ is labelled and *Q*_*in*_, the flow into the reservoir is indicated by the large arrow directed into the system. The termini linking to the microcirculation are the smallest vessel and the outflow *Q*_*out*_ is indicated by arrows going out of the system. The microcirculation (through which the reservoir discharges) and venous system are not shown as they are not considered part of the arterial reservoir.

The conservation of mass for the arterial network constrains the rate of change of the total volume, *V*, of the system to be equal to the difference between volume flow rate into the root, *Q*_*in*_, and the sum of the flow out of the terminal vessels, *Q_out_*:

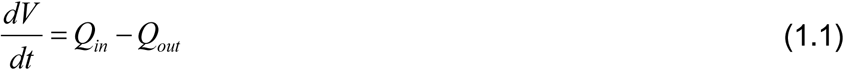

We assume that the end of each terminal vessel is coupled to a resistance,*R*_*k*_, which is assumed to be constant (i.e. independent of pressure). Under these conditions the flow out of the *k*^*th*^ terminal vessel is given by:

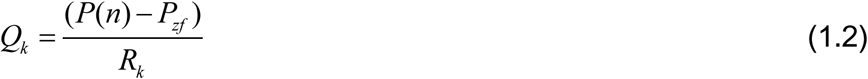

where *P*_*zf*_ (zero-flow pressure/critical closing pressure) is the pressure at which flow through the microcirculation ceases. As will be discussed later, *P*_*zf*_ can be greater than zero (or venous pressure), and for the purpose of this model it is assumed that it is the same for all termini. We also assume that the compliance of the *n*^*th*^ vessel is *C*_*n*_, where 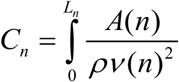 and *L*_*n*_, *A*_*n*_ and *v*_*n*_ are the length, cross-sectional area and wave speed respectively of the *n*^*th*^ vessel. The mass conservation equation can now be written in terms of the properties of the individual vessels

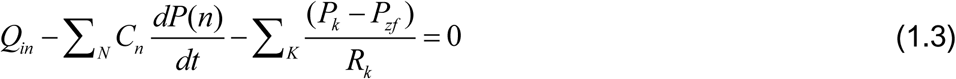

Where ∑_*N*_ is the sum over all of the vessels and ∑_*K*_ is the sum over the terminal vessels.

### Windkessel pressure

Frank’s 2-element Windkessel has similarities and differences from reservoir pressure. The Windkessel pressure *P*_*wk*_ is assumed to be uniform.(5) With this assumption, the pressure, *P*_*wk*_ can be taken outside of the summations and the mass conservation equation reduces to:

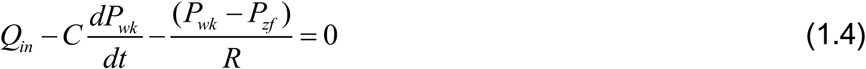

where the total arterial compliance, *C* = ∑_*N*_*C*_*n*_ and the total peripheral resistance, 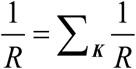, using the usual formula for resistances in parallel.

This is, in essence, the approach originally used by Frank to solve for the Windkessel pressure, although Frank assumed that *P*_*zf*_ *= 0*.

*P*_*wk*_ takes account of the compliant nature of the large arteries but the assumption of a uniform pressure implies an infinite wave speed, which is physiologically implausible. The modified reservoir pressure does not share this defect.

### Reservoir pressure and excess pressure

The modified reservoir pressure is defined as a pressure that is similar in form throughout the extent of the arterial reservoir, but which is delayed by the time it takes for waves to travel from the root to that location; hence, the reservoir pressure in the *n*^*th*^ vessel is

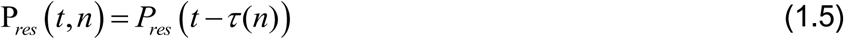

where, *τ(n)*, the wave transit time, is the time it takes for waves to travel from the root to vessel *n*. With this assumption, the mass conservation equation takes the form:

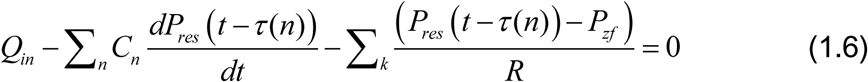

This equation involves only one pressure, *P*_*res*_, instead of involving *N* different pressures, *P(n)*. This equation is a first-order time-delay differential equation with constant coefficients. These have been studied extensively in the context of control theory and there are existence and uniqueness theorems that ensure that a solution of this equation exists, with suitable boundary conditions, and that it is unique. Unfortunately, there is no established way to find the solution for a particular case, and most solutions are found by iterative methods. Solving the equation would require knowledge of all of the individual compliances and resistances of all of the arteries - knowledge that is impossible to obtain in practice since there are too many vessels.

The excess pressure (*P*_*xs*_) in the *n*_*th*_ vessel is defined as the difference between the measured pressure and the reservoir pressure

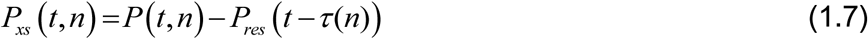

with these definitions *P*_*res*_ and *P*_*xs*_ can be calculated as shown below.

### Calculation of reservoir pressure

All of the methods of estimating the reservoir pressure are based on the assumption that the wave transit times are small in comparison with the cardiac cycle.(τ(n) ≪ 1 cardiac period – assumed to be ∼ in man). This is supported by *in vivo* measurements of the aorto-iliac transit time in humans (i.e. the time taken for the initial compression wave to traverse the whole of the aorta from aortic root to iliac bifurcation) which are <80 ms,(21) while the time from foot to peak pressure is approximately 2.5-fold longer (∼200 ms(22)).

So, using a Taylor expansion for *P*_*res*_*(t − τ(n))*,

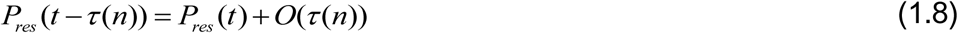

where *O(τ(n))* stands for terms of order, *(τ*_*n*_ Substituting into the mass conservation equation, the terms involving *P*_*res*_ can be taken out of the summations and we obtain the ordinary differential equation (ODE):

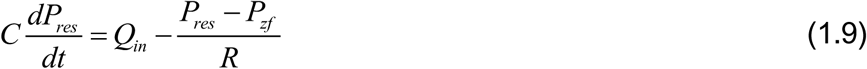

where, as in the derivation of the Windkessel pressure,*C* = ∑_*N*_*C*_*n*_, and 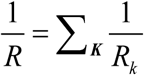. This shows that to *O(τ(n))*, the equation for *P*_*res*_ is identical to the equation for *P*_*wk*_, but without the need to assume a uniform pressure throughout the reservoir. This also makes clear that the reservoir pressure travels as waves, and is the basis of the method of estimation of *P*_*res*_.

### Calculating P_res_ when pressure and aortic flow are known

If the aortic inflow, *Q*_*in*_*(t)* is measured simultaneously with the pressure *P*_*0*_*(t)*, then the calculation of *P*_*res*_*(t)* is relatively straightforward. The solution of the ODE is easily found by quadrature

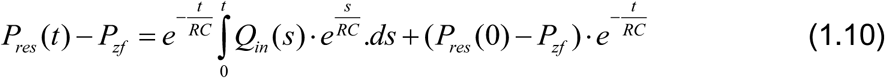

where *s* is time from the start of systole. During diastole when the valve is closed, *Q*_*in*_ *= 0*, and the solution becomes

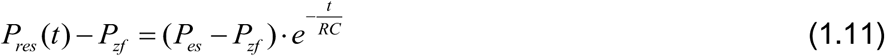

where *P*_*es*_ is the pressure at end-systole. This is a mono-exponentially falling function of time with a time constant, *τ* = *RC*. This is one of several well-established ways to estimate total arterial compliance,(23) although it should be noted that inclusion (or not) of *P*_*zf*_ has a substantial effect on estimates of the time constant or arterial compliance using this method.(24-26) We use maximum negative rate of pressure change (*max -dP/dt)* as the indicator of end-systole to determine *P*_*es*,_ since the timing of this event has been shown to agree very closely (mean error <0.4ms) with the time of cessation of aortic flow at the end of systole in invasive studies in dogs(27) and is easy to identify in recorded pressure waveforms.

Frequently reservoir pressure calculations are performed using flow velocity rather than volumetric flow rate (Q), under these circumstances it should be remembered that while estimates of *P*_*res*_, *P*_*zf*_ and *τ* are unaffected, the values of *R* and *C* will equal resistance ⨯ area and compliance/area, where the area refers to the cross sectional area of the aorta.

This method of calculating *P*_*res*_ can be used in experiments where both pressure and flow rate (or velocity) are measured in the aorta. Clinically, however, it can be difficult to obtain simultaneous measurements of pressure and flow and so another more approximate method has been devised for calculating *P*_*res*_ that requires only the pressure to be measured.

### Calculating P_res_ when only pressure is known

The method for calculating *P*_*res*_ using only pressure measurement is based on an observation made by Wang et al.(7) in dogs, who reported that the excess pressure, *P*_*xs*_ was directly proportional to the flow into the aortic root, *Q*_*in*_. Subsequent studies in humans employing invasive measurements of pressure and flow velocity in the aorta(28) and non-invasive measurements of carotid artery pressure and aortic flow(29, 30) have made similar observations. On this assumption, we can substitute *Q*_*in*_ *=*ζ*P*_*xs*_ *=* ζ*(P-P*_*res*_) into the mass conservation equation, where ζ is a constant of proportionality. If this relationship is viewed as analogous to a 3-element Windkessel,(31) then ζ will be related to the characteristic admittance, or *1/Z*_*c*_, (i.e. the inverse of the characteristic impedance).(29) Indeed, Westerhof and Westerhof(15) have proposed that if the analogy with the 3-element Windkessel holds, then *P*_*res*_ will be equal to twice the backward pressure (*P*_*b*_). (This relationship can be shown to be true in diastole when aortic flow (*Q*) = 0, but the derivation relies on the assumption that *Q = Q*_*in*_ which may be questionable.)

If we define *k*_*s*_ *=* ζ*/C* and *k*_*d*_ *= 1/RC*, then equation (1.9) can be written as:

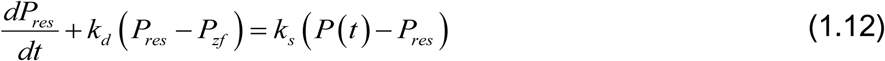

This equation is similar in form to the previous ODE, but the right-hand side depends on *P(t)* rather than *Q*_*in*_.

This first-order linear differential equation can be solved as:

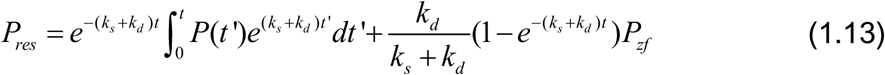

This equation can be solved by iterative non-linear regression based on a 3-element Windkessel model,(32) or alternatively the diastolic parameters *k*_*d*_ and *P*_*zf*_ can be estimated by fitting an exponential curve to the pressure during diastole,

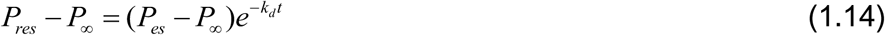

where the offset of the fit (*P∞*) is assumed to be equal to *P*_*zf*_ (NB the validity of this assumption is examined below). Then *k*_*s*_ is estimated by minimising the square error between *P* and *P*_*res*_ obtained over diastole in equation 1.12.

This formulation of reservoir and excess pressure makes clear the difference between Windkessel and reservoir pressure as previous noted by Alastruey(33) (Figure 2).

**Figure 2.**
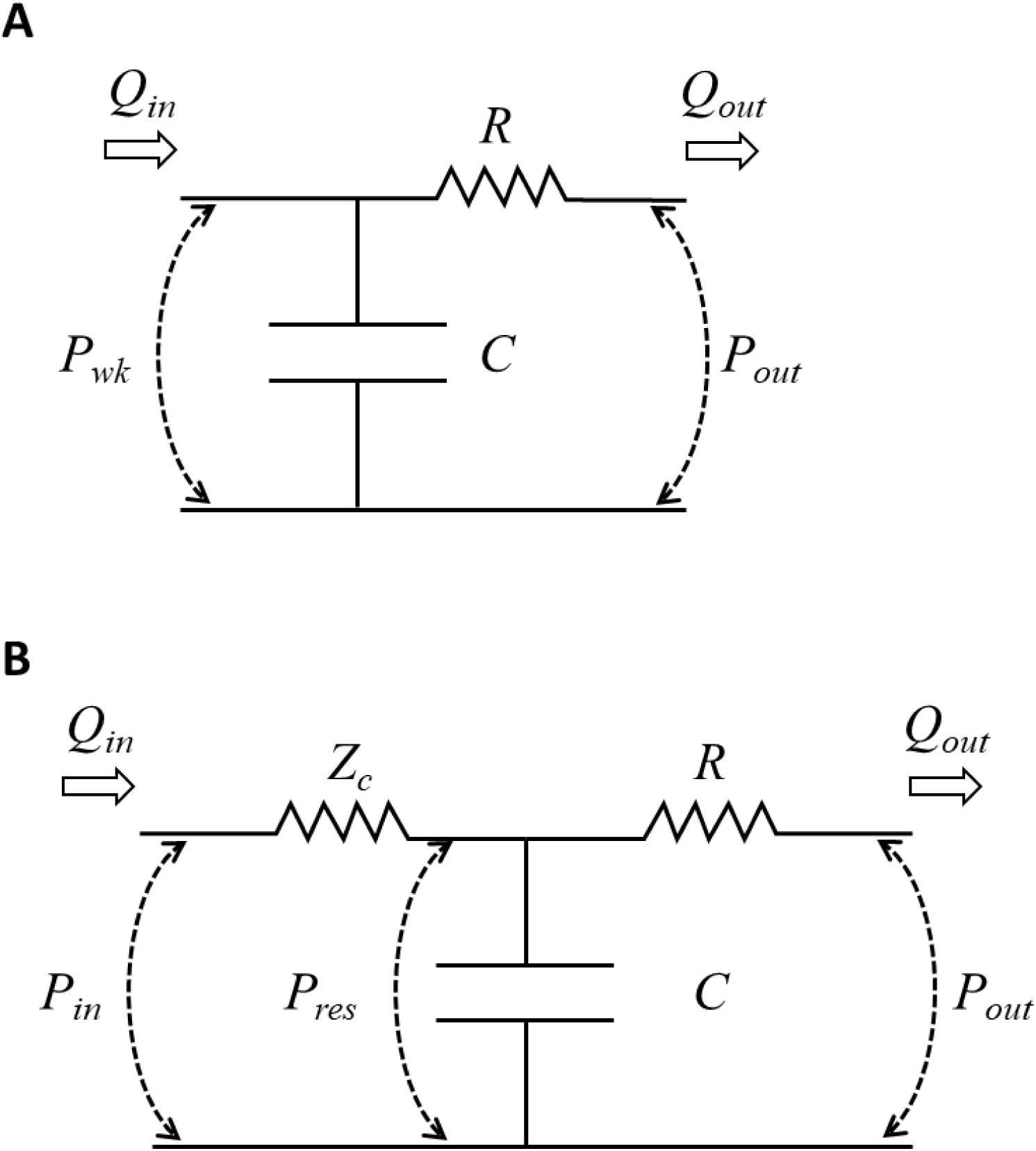
Circuit diagrams illustrating the comparison between a 2-element Windkessel and reservoir pressure conceptualised as a 3-element Windkessel model. A) 2-element Windkessel; here *P*_*in*_ is equal to Windkessel pressure (*P*_*wk*_) B) 3-element Windkessel model of reservoir pressure (*P*_*res*_). In the 3-element model, *P*_*res*_ still corresponds to the pressure across the capacitance, *C*, but *P*_*res*_ is smaller than *P*_*in*_ due to the pressure drop across the characteristic impedance, *Z*_*c*_. Modified from Alastruey(33).

In principle the approach described here should only be valid if *Q*_*in*_ *=* ζ*P*_*xs*_, which implies the absence of reflections. As discussed, reflections are always present in the circulation, but this assumption may hold within reasonable limits in the proximal aorta of healthy individuals (7, 34, 35). It is less likely to be true in more peripheral locations where prominent wave reflections are observed in early systole.(36, 37)

The assumption of proportionality between *Q*_*in*_ and *P*_*xs*_ may also not apply when there is pathology giving rise to marked wave reflections in the proximal aorta.(38, 39) Despite these provisos, estimates of *P*_*res*_ made at various locations in the aorta (the transverse aortic arch, the diaphragmatic aorta, the aorta at the level of the renal arteries, and the aortic bifurcation)(40), and in the brachial and radial artery using invasive methods(41) are very similar to estimates in the proximal aorta (within 5%). Estimates of *P*_*res*_ made using non-invasive methods also show acceptable concordance with aortic measures (intra-class correlation coefficient 0.77), although they are less accurate, probably due to errors in the estimation of systolic and diastolic pressure by cuff methods.(42) In contrast, as expected, estimates of *P*_*xs*_ differ substantially between the proximal aorta, and more peripheral locations,(40-42) with *P*_*xs*_ being larger in more proximal locations. Alastruey(33) proposed an alternative definition of *P*_*xs*_ where it would be re-defined as proportional to flow at any location, however such a proposal would also result in a redefinition of *P*_*res*_ and so far the value of this approach seems not to have been explored.

Currently, there appear to be no publications where estimates of *P*_*res*_ based on pressure and flow velocity have been compared with estimates derived from pressure alone, although unpublished data from our group indicates excellent agreement between the approaches (correlation coefficient >0.9; mean difference in peak *P*_*res*_ = 2±1 mm Hg, p<0.002) based on invasive measurements of pressure and flow velocity in the aorta.

## The relation of *P*_*zf*_ to *P*_*∞*_ and the physiological interpretation of *P*_*zf*_

The presence of positive pressure in the arterial circulation following cessation of flow, *P*_*zf*_, has been recognised for many years.(43) In the context of the arterial reservoir, some authors have assumed that *P*_*zf*_ corresponds to venous pressure (or zero as a rough approximation to venous pressure),(29, 44) or else that it should represent mean circulatory filling pressure (MCFP).(32, 45) MCFP, as defined by Guyton is ‘the pressure that would be measured at all points in the entire circulatory system if the heart were stopped suddenly and the blood were redistributed instantaneously in such a manner that all pressures were equal.’(quoted in (46)).

Previous work in several species (47, 48) including some necessarily limited work in man(49) has shown that *P*_*zf*_ differs from venous pressure and that in most(47, 48, 50, 51) (but not all cases(52)) there is no equalization of arterial pressure with venous or right atrial pressure, even after prolonged cessation of flow. Whether *P*_*zf*_ corresponds to MCFP has not been formally examined previously as far as we can tell, so in order to address this question we undertook a systematic review of the literature; some of these data have been published previously in abstract form.(53)

A literature search was performed using PubMed and was limited to full articles in English using the search terms “mean circulatory filling pressure” OR “Mean systemic filling pressure” OR “critical closing” OR “zero-flow” in publications prior to 01/09/2019. Only data relating to measurements of pressure following cessation of systemic flow were included; other exclusions were: individual case-reports, pregnancy, non-adult animals, not mammalian, post-mortem, or any non-human models of disease. Meta-analysis was performed using a random effects model since it was anticipated that there would be heterogeneity between studies. Analyses were conducted in Stata 15.1. Data are shown as means (95% confidence intervals).

A total of 1255 unique publications were identified after removal of duplicates; 1235 were excluded during screening. The remaining 20 studies (48-51, 54-69) with *P*_*zf*_ data were included in a meta-analysis (**Figure 3**); these included data from dog, rat, pig and human; 8 of these articles also provided data on MCFP from the same studies. Some further details of these studies are shown in **Supplementary Table S1**. From this analysis *P*_*zf*_ = 26.5 (23.4, 29.5) mmHg (**Figure 4**; 20 studies; mean(95% confidence interval); n = 311; I^2^ = 97%; P <0.001) and MCFP = 10.6 (9.3, 12.0) mmHg (8 studies; n = 178; I^2^ = 96%; p < 0.001). The difference between *P*_*zf*_ and MCFP was 15.1 (12.0, 18.3) mmHg (8 studies; n = 178; I^2^ = 97%; p < 0.001). The comparison between *P*_*zf*_ and MCFP is shown graphically in **Figure 5**. There was no evidence of small sample bias based on an Egger test for either analysis (p>0.05 for both). Further analyses provided no convincing evidence that the duration of cessation of flow was related to the estimate of *P*_*zf*_ based on meta-regression (p = 0.1) although the small sample size precluded firm conclusions. Similarly, the extent of heterogeneity within subgroups (e.g. species, method of calculation) prevented any reliable conclusions on the importance of these factors in the observed heterogeneity in *P*_*zf*_ between studies. Nevertheless it seems plausible that methodological differences between studies contribute to variability in the estimates of *P*_*zf*_. There was evidence that MCFP differed between species (test for heterogeneity between sub-groups p = 0.007) but there was insufficient data to examine whether reported differences between studies contributed to heterogeneity in MCFP.

**Figure 3.**
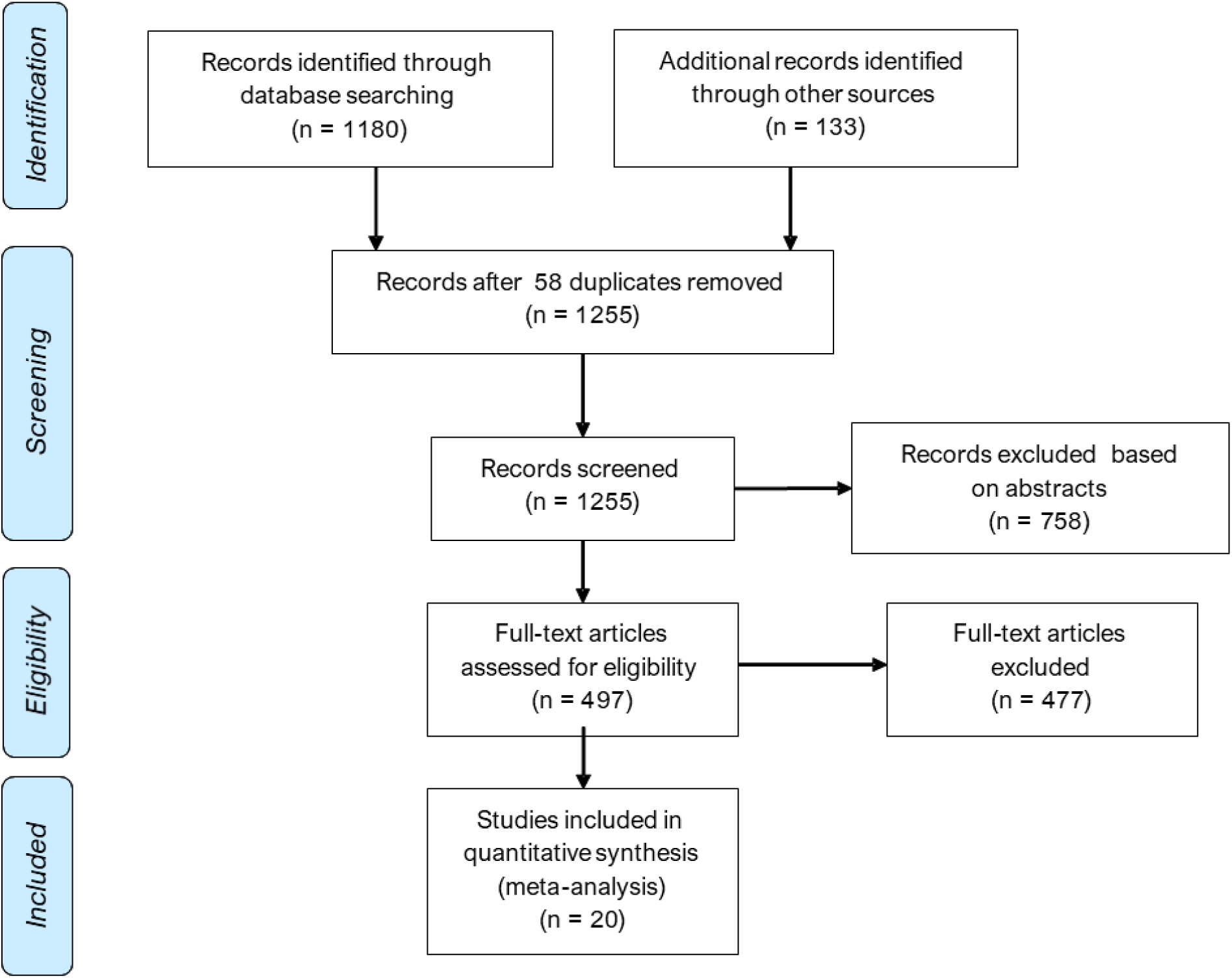
A Preferred Reporting Items for Systematic Reviews and Meta-Analyses (PRISMA) flow diagram for zero-flow pressure (*P*_*zf*_).

**Figure 4.**
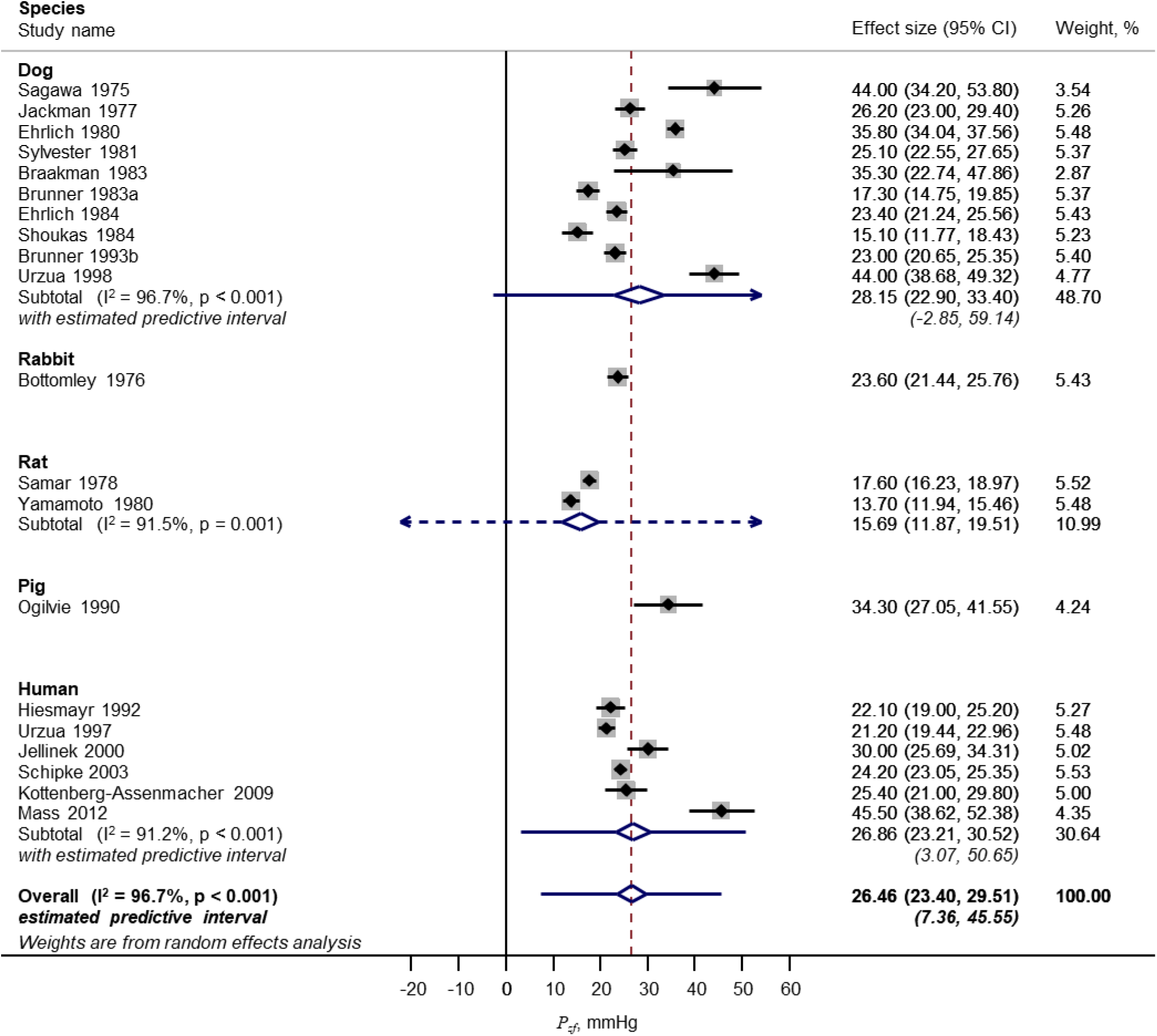
Forest plot of meta-analysis of zero-flow pressure (Pzf). Data categorised by species.

**Figure 5.**
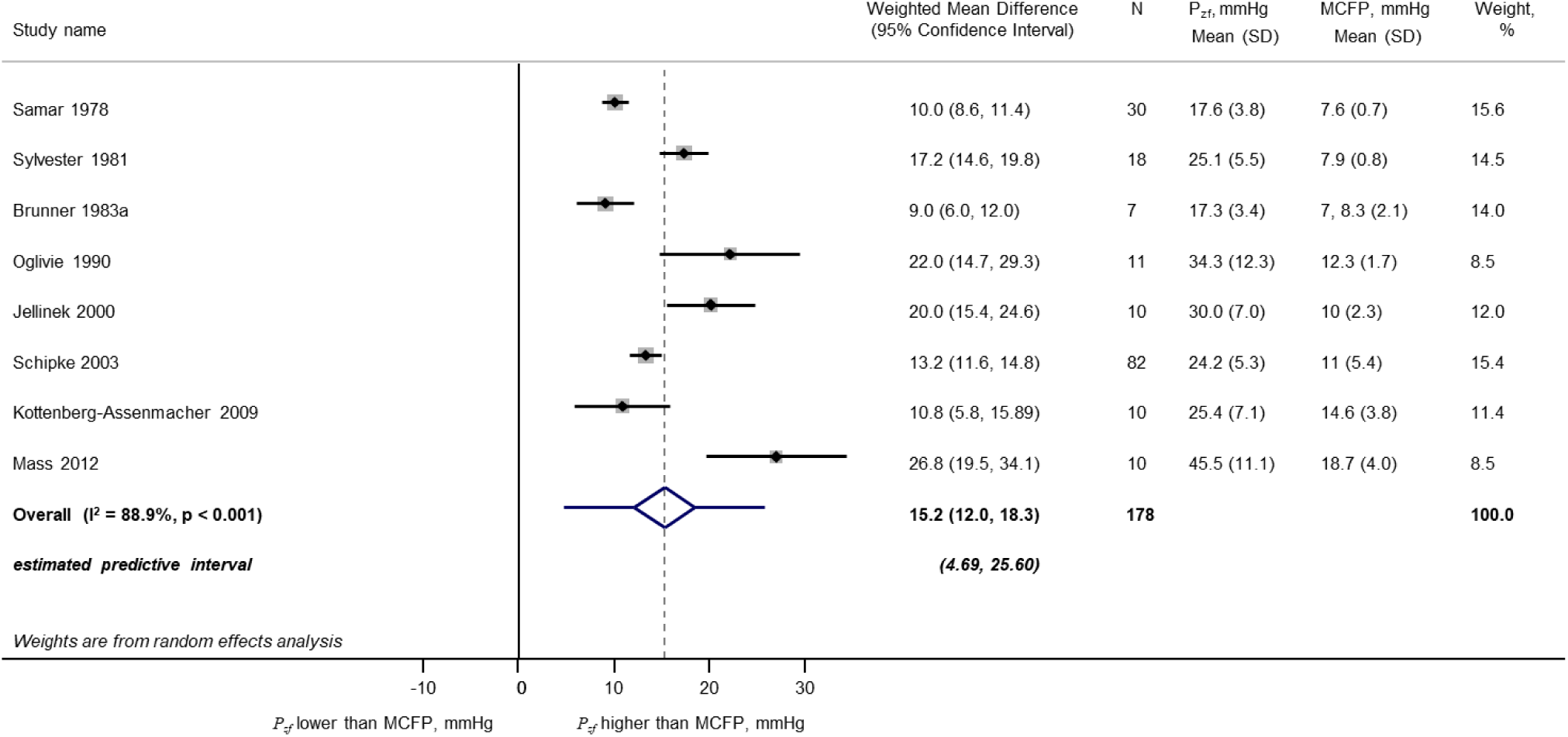
Forest plot of meta-analysis of differences between zero-flow pressure (*P*_*zf*_) and mean circulation filling pressure (MCFP) in studies in which both were measured.

Based on these data it seems clear that despite considerable heterogeneity *P*_*zf*_ does not equal MCFP. It is noteworthy that this is consistent with the standard practice in many experiments designed to estimate MCFP which either routinely transfer blood from the arterial to the venous circulation to achieve equilibration of pressure,(57, 70) or else apply a correction factor to take account of the ‘trapped’ volume of blood in the arterial circulation after cessation of flow.(57) We note that this finding does not necessarily imply that arterial and venous pressures cannot equilibrate after extremely prolonged cessation of flow.(71) The duration of cessation of inflow in the studies identified in the systematic review was between 3 and 30s (median 12.5s), which is longer than the time constant of the decline in pressure (typically ∼2 to 3s) but short enough to at least partially limit the secondary rises in MCFP due to reflex changes vasomotor tone and decreased venous compliance that tend to reduce differences between MCFP and *P*_*zf*_ through elevated MCFP. (71) It is also unlikely that substantial oedema or hemodilution would occur over this time span and confound estimates of MCFP and *P*_*zf*_. Most studies did not measure flow as well as pressure, but those that did reported that cessation of flow occurred between 3 to 20s after cardiac arrest or switching off the perfusion pump.(56, 58, 62) This suggests that the duration of cessation of inflow was probably sufficient to obtain a reliable estimate of *P*_*zf*_.

Values of *P*_*zf*_ that exceed MCFP are consistent with previous suggestions that *P*_*zf*_ represents a pressure due to a Starling resistor effect, sometimes termed (although some argue inappropriately(72, 73)) a ‘vascular waterfall’.(74) *P*_*zf*_ is also often termed critical closing pressure after Burton,(75) although it is now generally accepted that vessel closure does not account for *P*_*zf*_.(76-78) *P*_*zf*_ is not a fixed parameter and varies between species,(14) individuals,(79) physiological (or pathophysiological) conditions,(79) tissues(58, 61, 80) or even within tissues.(81) Given the reported between-tissue differences in *P*_*zf*_ it is likely that systemic *P*_*zf*_ is a weighted average of multiple *P*_*zf*_. This may relate to differences in vascular resistance between tissues since there is evidence that vasoconstriction and vasodilation respectively increase and decrease *P*_*zf*_ quite markedly.(14, 54, 82) Indeed in the rabbit ear vasoconstriction has been reported to increase *P*_*zf*_ to ∼130 mmHg — in excess of mean arterial pressure in the rabbit!(14) Increased tissue interstitial pressure also influences *P*_*zf*_, as would be expected from a Starling resistor.(83) Another factor that influences *P*_*zf*_ is blood rheology; estimates of *P*_*zf*_ are lower following hemodilution(48), although since positive *P*_*zf*_ has been observed using physiological saline,(43, 84) it seems unlikely blood-related factors such as red cell aggregation,(85) the complex viscous behaviour of blood at low flow,(86) or leukocyte plugging(87) fully account for *P*_*zf*_.

The duration of cessation of flow has been reported to affect the estimated *P*_*zf*_. (88) Short duration cessation of flow may result in over-estimates due to the effects of capacitive discharge from downstream vessels as suggested by Spaan(89) and Magder(90). This effect, which assuming a simple *RC* compartment model increases ‘apparent *P*_*zf*_ ‘ by an amount equal to *-*¢*RdP/dt* (where ¢ is the downstream microvascular compliance),(91) will introduce a difference between *P*_*∞*_ and *P*_*zf*_ that depends on the rate of pressure decline in diastole. Estimates of the effect of capacitive discharge have been made in the coronary circulation and show that this effect could easily account for a ∼10mmHg difference between *P*_*∞*_ and *P*_*zf*_.(91) Alternatively longer periods of flow cessation may lead to changes in vascular properties due to lack of flow or ischaemia or alterations in rheology or tissue interstitial pressure. Braakman et al.(82) presented evidence for two *P*_*zf*_ (instantaneous [arteriolar] and steady-state [venous]) in skeletal muscle, with the ‘instantaneous’ *P*_*zf*_ being dependent on vasoconstrictor tone whereas the ‘steady-state’ *P*_*zf*_ was not. It seems likely that the different techniques used to estimate *P*_*zf*_ will be differentially influenced by the factors that influence *P*_*zf*_ and that this will contribute to variation in estimates of *P*_*zf*_.

Despite these considerations, evidence presented here suggests that *P*_*zf*_ is substantially lower than estimates of *P*_*∞*_ (based on fitting diastolic pressure to equation 1.14). *P*_*∞*_ in the human aorta has been reported to be between 54 to 75 mmHg,(26, 40) while the upper limit of the predictive interval of *P*_*zf*_ in humans in our meta-analysis was 51mmHg. This is consistent with a previous report in humans where *P*_*∞*_ calculated from normal beats was ∼29 mmHg greater than *P*_*zf*_ calculated during arrest.(49) Estimates of *P*_*∞*_ will include uncertainties associated with fitting only short durations of diastole, particularly if the fit includes the perturbation in pressure that accompanies the onset of isovolumic contraction prior to the foot of the next pressure cycle.(92) However, this seems insufficient to account for such a large difference and it may indicate that one or more assumptions in the present approach to fitting reservoir pressure is not valid.

The assumption of a mono-exponential decline in diastolic pressure has been examined experimentally by a few authors. In 7 patients in whom aortic pressure was measured invasively, Liu et al(24) looked at whether estimates of the slope of the semi-log regression of pressure and time gave consistent estimates of *τ*, and whether regression of *dP/dt* versus *P* was linear during diastole; they found that results using either method were inconsistent with a mono-exponential decline. Kottenberg-Assenmacher et al.(49) reported that the goodness of fit (by *χ*^2^) of the time-dependent decline in invasive aortic pressure following circulatory arrest in humans was slightly better using a two-exponential model, or using a model including a pressure-dependent coefficient, although the effect on the estimates of these more complex models on the estimates of *P*_*∞*_ was small (<2mmHg). Schipke et al.(51) reported that the correlation coefficients (presumably to the linearized semi-log transformation of the mono-exponential function) were 0.92±0.05, and that data were less well fitted by linear and quadratic functions, but other functions seem not to have been examined. Brunner et al.(48) reported that after stopped flow the relationship between the natural logarithm of the declining pressure with time was linear in seven out of thirteen dogs (consistent with a mono-exponential decline) but in the remaining six dogs the data were not consistent with a mono-exponential decline. Sylvester et al.(50) reported that the decline in pressure following stopped flow was ‘well-described’ by a mono-exponential function with standard deviations on average <2mmHg but provided no other quantification of fits.

On the basis of theoretical considerations regarding small artery compliance,(89, 90) the pressure dependence of large artery compliance(24, 93) and resistance, (89, 94) it seems unlikely that the assumption of a mono-exponential decline should be valid.(89, 90) Still, in view of the well-recognised difficulty in fitting multiple exponentials to complex data,(95, 96) we believe that the fitting of multi-exponential functions to diastolic pressure is unlikely to yield much advantage, although it may be worth exploring.

## Conclusions

The arterial Windkessel is undoubtedly a simple and widely used conceptual model of the circulation. In its modified form the arterial reservoir can be viewed as analogous to the Windkessel but comprising multiple reflected (and re-reflected) waves that arise due to the different forward and backwards impedance properties of a branching network,(20, 97, 98) which give rise to a “horizon effect”.(16) The waves that make up the reservoir are indiscernible by wave intensity analysis as their individual magnitudes are so small,(37) but together they make up a large store of energy. This energy, which is effectively trapped within the large elastic (conduit) arteries due to reflection and re-reflection provides the motive force for tissue perfusion during diastole. This wave entrapment (45) equates to the volume storage of a classic Windkessel and accounts for the apparent similarities between these models. The waves that make up the reservoir persist across several cardiac cycles and account for most of the energy present in any particular cycle at quasi-steady state.(35) We have previously proposed that mean arterial pressure should be viewed as largely a product of these waves rather than the equilibrium state of the circulation as envisaged by Fourier-based impedance analysis.(99)

Following cessation of ejection, pressure declines in a quasi-exponential manner towards a value, *P*_*zf*_, which is the pressure at which outflow through the microcirculation ceases. A review of the experimental evidence suggests that *P*_*zf*_ exceeds venous pressure or MCFP, but also that estimates of the offset (*P*_*inf*_) derived from fitting a mono-exponential function to the decline in pressure during diastole of a normal cardiac cycle are substantially greater than *P*_*zf*_, possibly as a consequence of the mono-exponential assumption. Further work is required to establish on how best to estimate *P*_*zf*_ from recordings of normal cardiac cycles.

Several features of the modified arterial reservoir model (and its underlying wave nature) contrast with interpretations that would be made if the circulation were viewed as analogous to a single tube, and the utility of the single tube model is questionable in our view and that of others.(100) While tangential to the content of this review it is worth noting that the presence of multiple re-reflections also casts doubt on the utility of the ratio of forward to backward pressure, Pb/Pf (often termed reflection magnitude) as a measure of reflection since a substantial part of forward pressure will arise from re-reflection of initially reflected (backward travelling) waves. This issue has also been alluded to previously,(45, 100-102) but its implications for pulse wave analysis seem not to have been fully apprehended.

In summary the modified arterial reservoir represents a useful, albeit reduced, model of the circulation. The ability of parameters derived from this model to predict future cardiovascular events independent of conventional cardiovascular risk factors(103-111) suggests this model has clinical utility. No model of the circulation is perfect however, and its limitations need to be recognised.

## Data Availability

The data that support the findings of this study are available from the corresponding author, [ADH], upon reasonable request.

## Acknowledgements

AH receives support from the British Heart Foundation (CS/13/1/30327, PG/13/6/29934, PG/15/75/31748, CS/15/6/31468, PG/17/90/33415, IG/18/5/33958), the National Institute for Health Research University College London Hospitals Biomedical Research Centre, the UK Medical Research Council (MR/P023444/1) and works in a unit that receives support from the UK Medical Research Council (MC_UU_12019/1).

